# Clinical characterization of respiratory droplet production during common airway procedures using high-speed imaging

**DOI:** 10.1101/2020.07.01.20144386

**Authors:** SK Mueller, R Veltrup, B Jakubaß, JS Kempfle, S Kniesburges, MJ Huebner, H Iro, M Döllinger

**Author notes:** **Corresponding author:** Sarina K. Mueller, MD, Department of Otolaryngology, Head and Neck Surgery, University Hospital Erlangen, Friedrich-Alexander-Universität Erlangen-Nürnberg (FAU), Waldstrasse 1, 91054 Erlangen. **Declarations:**. **Financial disclosures:** This research did not receive any specific grant from funding agencies in the public, commercial, or not-for-profit sector. **Authors’ contributions:** SKM, RV, MJH performed the experiments. SKM, RV, BJ, SK, MD, JSK and HI analyzed the data. SKM drafted the manuscript. All authors edited the manuscript.

## Abstract

**Background:** During the COVID-19 pandemic, a significant number of healthcare workers have been infected with SARS-CoV-2. However, there remains little knowledge regarding droplet dissemination during airway management procedures in real life settings.

**Methods:** 12 different airway management procedures were investigated during routine clinical care. A high-speed video camera (1000 frames/second) was for imaging. Quantitative droplet characteristics as size, distance traveled, and velocity were computed.

**Results:** Droplets were detected in 8/12 procedures. The droplet trajectories could be divided into two distinctive patterns (type 1/2). Type 1 represented a ballistic trajectory with higher speed droplets whereas type 2 represented a random trajectory of slower particles that persisted longer in air. Speaking and coughing lead to a larger amount of droplets than non-invasive ventilation therapy. The use of tracheal cannula filters reduced the amount of droplets.

**Conclusions:** Respiratory droplet patterns generated during airway management procedures follow two distinctive trajectories based on the influence of aerodynamic forces. Speaking and coughing produce more droplets than non-invasive ventilation therapy confirming these behaviors as exposure risks. Even large droplets may exhibit patterns resembling the fluid dynamics smaller airborne aerosols that follow the airflow convectively and may place the healthcare provider at risk.

## Background

The severe acute respiratory syndrome coronavirus 2 (SARS-CoV-2) leads to a high variability of symptoms. While some exhibit minor symptoms of an upper respiratory infection, some patients presents with severe sequelae similar to the acute respiratory distress syndrome (ARDS)^1–3^. With worldwide dissemination of the disease and the increasing number of deaths, the modes of transmission in the healthcare environment is a subject of significant investigation ^4,5^. Many medical providers have been infected during the pandemic which in some cases has lead to death. It is therefore paramount to minimize healthcare worker exposure during highly “aerosolizing” procedures^6,7^. Multiple recommendations to ensure maximal safety for the medical personnel during airway management procedures are being developed and regularly updated^6–11^. This includes limiting some procedures to the minimum necessary, including non-invasive ventilation or tracheal cannula changes in SARS-CoV2 positive patients. Generally, aerosol transmission is described to occur by droplets (diameter > 5 µm)^3,8,12^ and airborne aerosols/ droplet nuclei (diameter <5 µm). However, precondition for an aerosol transmission is that the virus retains infectivity and replicability within these small particles^12^. For SARS-CoV-2, there are already studies that have identified viable virus within aerosols up to hours^13,14^. The number of detectable viruses following coughing versus exhaled breath is currently under investigation^15,16^.

While the current consensus presumes primary transmission through respiratory droplets^4,5,12,17^, there remains no quantitative data concerning the droplet trajectories and fluid patterns during airway management procedures in real life settings. In order to estimate the risk to medical personnel and to develop appropriate safety precautions it is important to analyze droplet size, aerodynamic characteristics and trajectories during different procedures. Therefore, the objective of this study was to assess the frequency, size and velocity of large respiratory droplets as well as characterize aerodynamic droplet patterns during common airway management procedures using high-speed imaging. The airway management procedures analyzed included non-invasive CPAP (continuous positive airway pressure) ventilation, high flow ventilation, tracheotomy cannula change, extubation, nebulizing and SARS-CoV-2 swab as well as common behavioral patterns including speaking and coughing.

## Methods

### Study design and inclusion of patients

All experiments were conducted at the Friedrich-Alexander-University Erlangen-Nürnberg. All experimental protocols as well as the study design were approved by the ethics committee of the Friedrich-Alexander-University Erlangen-Nürnberg (No 167_20B). Informed consent was obtained from all participants for participation in the study. 12 different airway management procedures were investigated. A total of n=8 patients (6 males, 2 females) were included. Depending on the procedure, the individual scenarios were performed up to 5 times. All patients received the airway management procedures as part of their routine clinical care.

### Equipment, experimental setup and outcome measure

The patients were positioned in a routine posture. This included a supine position for the extubation and a seated for all remaining procedures. A sterile clean room was used for the experiments.

Our first outcome measure was to display droplets originating from the patients. The second outcome measure was to display droplets originating from the examiner wearing a FFP3 plus a surgical mask. For both outcome measures, the number of droplets, size, trajectories, distance travelled, and velocity were quantified.

To detect the droplets in the air, we developed a setup with light against a black background to record the scattered light with a high-speed camera. Two Fenix TK-35 LED lights were used to illuminate the scene in front of the patient (500 lumens each). The room as well as patient and examiner were covered in black clothing as background in order to display the illuminated particles as richly as possible. A high-end industrial high-speed camera Vision Research Phantom v2511 recorded the procedures. The camera was aimed perpendicular to the longitudinal axis of the patient at the height of head at a distance of 1.5 meters. The recorded region was an area in front of the patient to represent the location where the provider would potentially stand during procedures, i.e. distance of up to 0.9 m in front of the patient. The videos were recorded at a spatial resolution of 1280×800 pixels yielding a minimal spatial resolution between 440 µm and 660 µm per pixel. This spatial limitation is caused by the choice of region (i.e. up to 0.9 m in horizontal direction to track the droplet trajectories) and the physical dimensions of the camera chip (5/3 inch). The recording rate was set to 1000 fps (frames per second) to accurately track the droplets. The camera control software PCC 2.6 was used. A reference image of a measuring rod was taken for subsequent metric computation of droplet trajectories.

### Experimental Conditions

The following common airway management procedures were studied:

1. Non-invasive CPAP with PEEP (positive end expiratory pressure) of 5mbar, 6mbar, 8mbar and 10mbar with and without coughing
2. Non-invasive CPAP with a PEEP of 5mbar and a Δp support (pressure support ventilation) of 10mbar, 15mbar and 20mbar with and without coughing
3. Non-invasive CPAP without leakage, with 50% leakage and with 80% leakage with and without coughing
4. Nasal oxygen via a nasal tube (Dahlhausen, Köln, Germany) at 2l/min, 4l/min, 6l/min, 8l/min and 10l/min with and without coughing
5. High-flow nasal oxygen at 15l/min without coughing
6. Nebulizing with an oxygen mask (Micro Mist® Nebulizer plus mask, Hudson RCI, Wayne, PA, USA)
7. Tracheal cannula suctioning with (7.1) and without (7.2) filter (hygroscopic condenser humidifier, Aqua+TS, Hudson RCI, Wayne, PA, USA)
8. Tracheal cannula removal without filter
9. Coughing and suctioning after tracheal cannula removal
10. Reinsertion of tracheal cannula without filter
11. Extubation (5.0 no cuff Vygon, Aachen, Germany)
12. SARS CoV-2 swab according to the WHO (World Health Organization) guidelines^18^.

For paradigm 1) to 3), a Dräger Evita® V800 respirator (Lübeck, Germany) was used. For paradigms 7) to 10), a Tracheoflex, 9,0 mm, with Cuff (Rüsch, Berlin, Germany) was applied. The nebulizing therapy was used for representation of fine particle aerosols (manufacturer’s information mass median aerodynamic diameter (MMAD) 3.6 µm).

Additionally, the paradigm speaking “stay healthy” was used as positive control (13).

### Image Processing and Quantification techniques

A software script specially designed for evaluating the recorded video data was implemented in MATLAB. It allows the manual tracking and computes the size of particles. Droplet sizes were found to be between 1 and 4 pixels. Based on the physical specification of the camera chip, particles were divided into three categories, large (1000 < d < 2000 µm), medium (670 µm < d ≤ 1000 µm) and small (d ≤ 670 µm). Traveled distances and velocity values of the particles were computed.

## Results

### Characterization of two main trajectory patterns

All visible droplets were tracked in each condition. No droplets were visible for paradigms 3, 5, 6, 11, 12. For paradigms 1, 2, 7.2, 8, and 9 all visible droplets were tracked. For paradigms 4, 7.1, 10 and 13 the number of droplets exceeded the ability for individual tracking. Comparing all scenarios, two major droplet trajectories were discerned based.

Type 1 represented a ballistic curve that descended in a predictable curve with a maximal velocity of 26.41 m/s. The velocity was maximal at start and exponentially decelerated over time to a minimal velocity of 0.04 m/s. The maximal distance travelled in horizontal x-direction was 0.73 m and in upper vertical y-direction 0.14 m. The average diameter of the droplets was 0.66 mm ± 0.32 mm. A paradigm with type 1 was e.g. 13) speaking “stay healthy” **(figure 1A**,**B)**.

**Figure 1:**
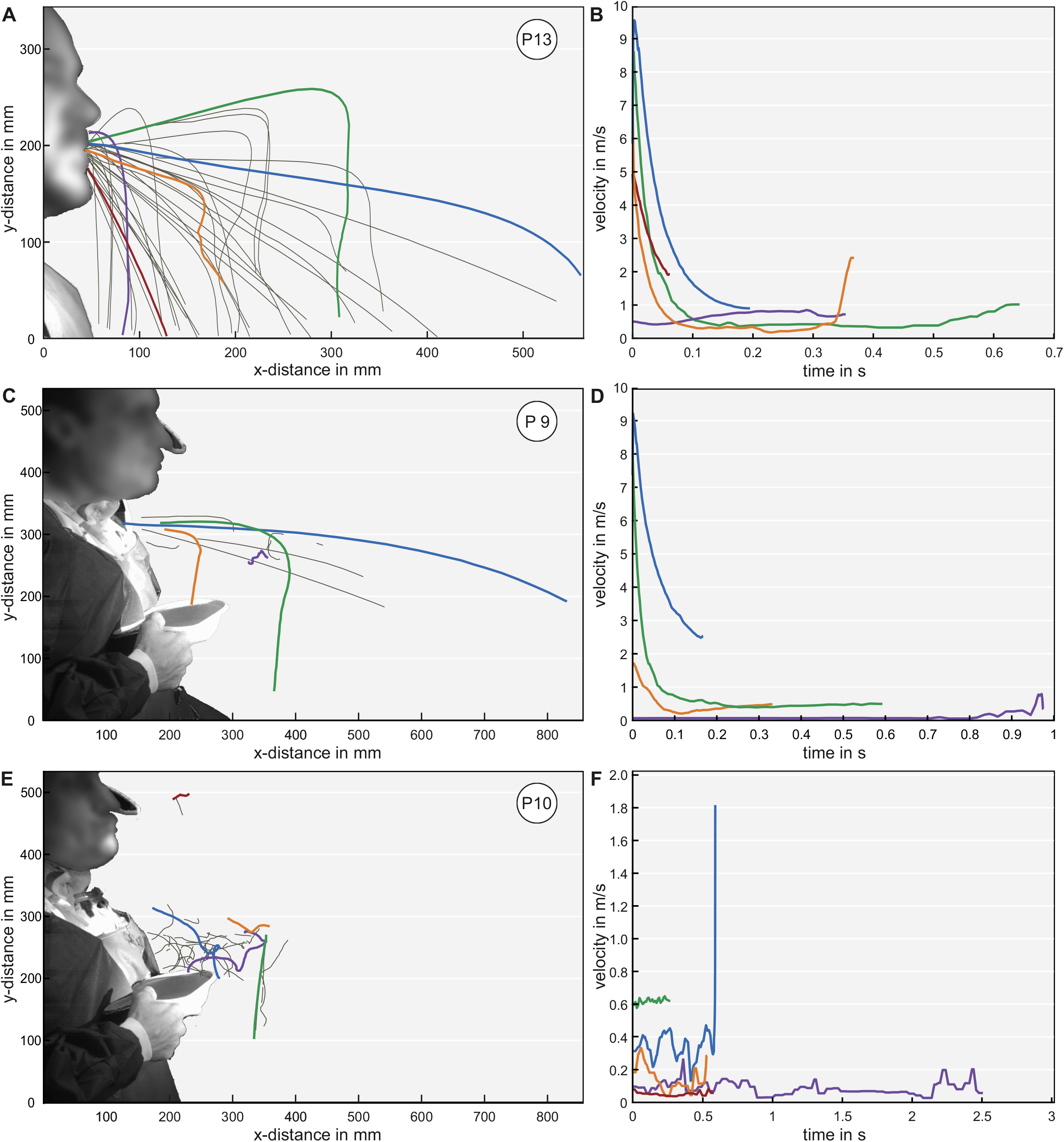
Examples of computed trajectories (left) and velocities (right) for tracked droplets within three conditions. The most representative trajectories were selected. The colored trajectories correspond to the matching velocity color: A) Droplet trajectories for the positive control speaking “stay healthy” (type 1 > type 2) B) Velocity-time diagram for speaking “stay healthy” C) Droplet trajectories for coughing out of the tracheostomy without tracheal cannula (type 2 > type 1) D) Velocity-time diagram for coughing out of the tracheostomy without tracheal cannula E) Droplet trajectories for the insertion of a tracheal cannula without a filter (type 2) F) Velocity-time diagram for the insertion of a tracheal cannula without filter.

Type 2 was unordered and non-directed. These droplets were mainly driven by convective flow within the ambient air with random acceleration and deceleration over time similar to the behavior expected by aerosols < 5 µm. The minimal velocity was 0.01 m/s and the maximal velocity was 7.02 m/s. The maximal distance travelled in horizontal direction was 0.70 m and 0.16 m in the upper vertical direction. Each droplet showed a different movement pattern. The average size of the droplets was again 0.66 mm ± 0.16 mm. A paradigm with type 2 was e.g. 10) reinsertion of the cannula **(figure 1C**,**D)**.

For all conditions that showed a type 1 pattern, there was also a type 2 pattern seen. This included all conditions (4, 7.1, 9, and 13) where coughing or speaking was performed **(figure 1 E**,**F; video 1)**. For paradigm 4, 7.1, 9, and 13, small to large droplet sizes were seen. Comparing the two patterns, type 2 droplets remained in the air after type 1 droplets already descended. However, for the conditions 1, 2, 7.2, 8 and 10 (non-invasive ventilation, tracheal cannula removal and reinsertion) only a type 2 pattern was seen. For conditions 1, 2, 7.2, and 10, only small droplet sizes were seen. For condition 8, there was also a large droplet size. For conditions 3, 5, 6, 11 and 12, no droplets could be seen **(figure 2A, B)**.

**Figure 2:**
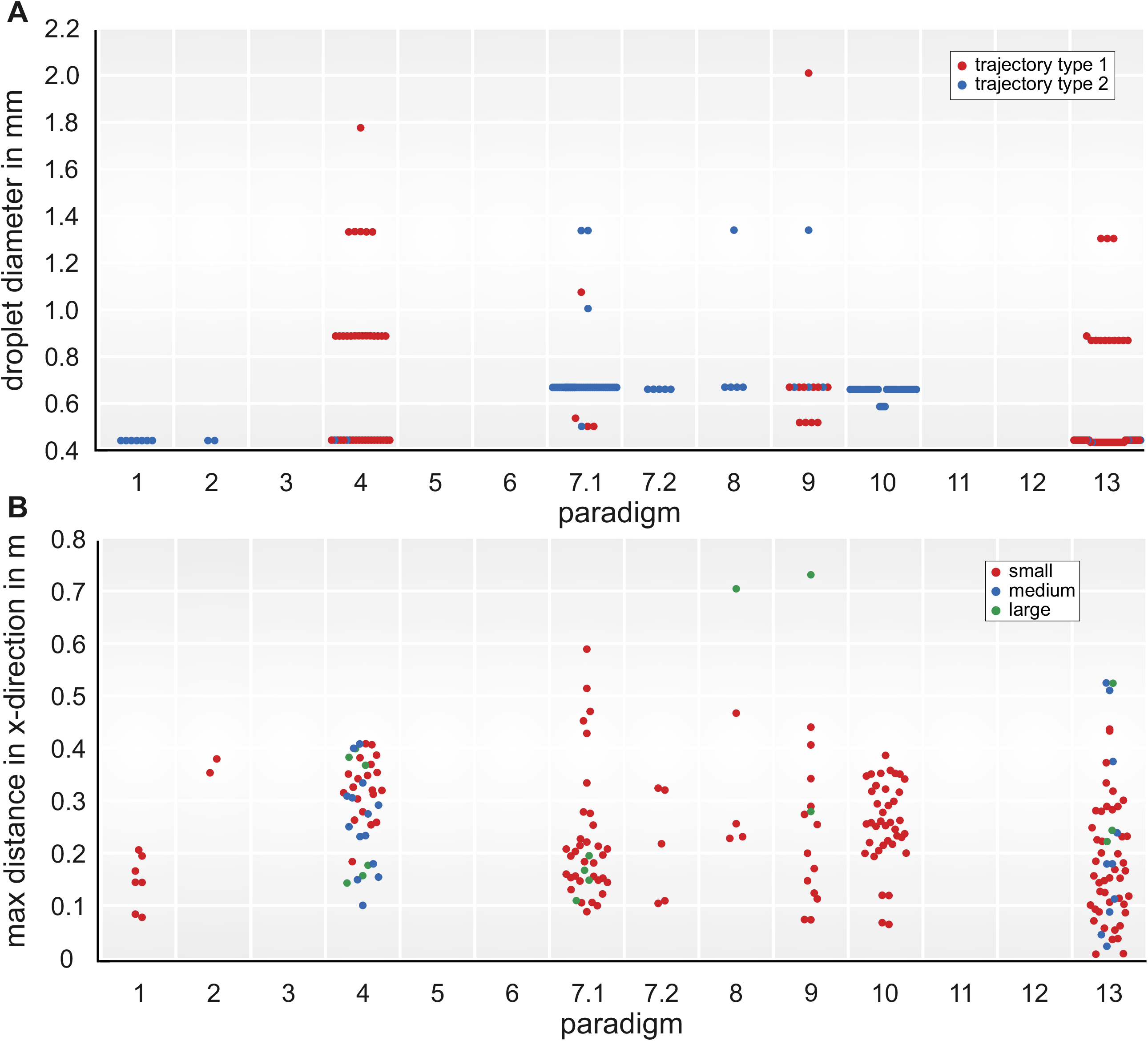
**A)** Beeswarm diagrams over all 13 paradigms representing (A) the droplet size separated for trajectory types and (B) the maximal distance travelled in horizontal × direction separated for the three droplet sizes. For A) the red color represents type 1 and the blue color type 2. For B) the red color represents small droplets, the blue color medium sized droplets and the green color large droplets.

### Reduction in droplet amount using a filter during tracheal cannula suctioning

Comparing tracheal cannula suctioning with and without filter, significant differences were seen. In the scenario without a filter (paradigm 7.1), there was a large amount of droplets with non-directed trajectories (type 2). With a filter (paradigm 7.2) there was an average 80% reduction in the amount of droplets with type 2 behavior **(figure 3 A**,**B)**. After removal of the filter, many droplets were visible and could be characterized as type 2 trajectories. In case of coughing after removal or insertion of the tracheal cannula, the droplet formation increased and changed to a combined type 1 and 2 pattern.

**Figure 3:**
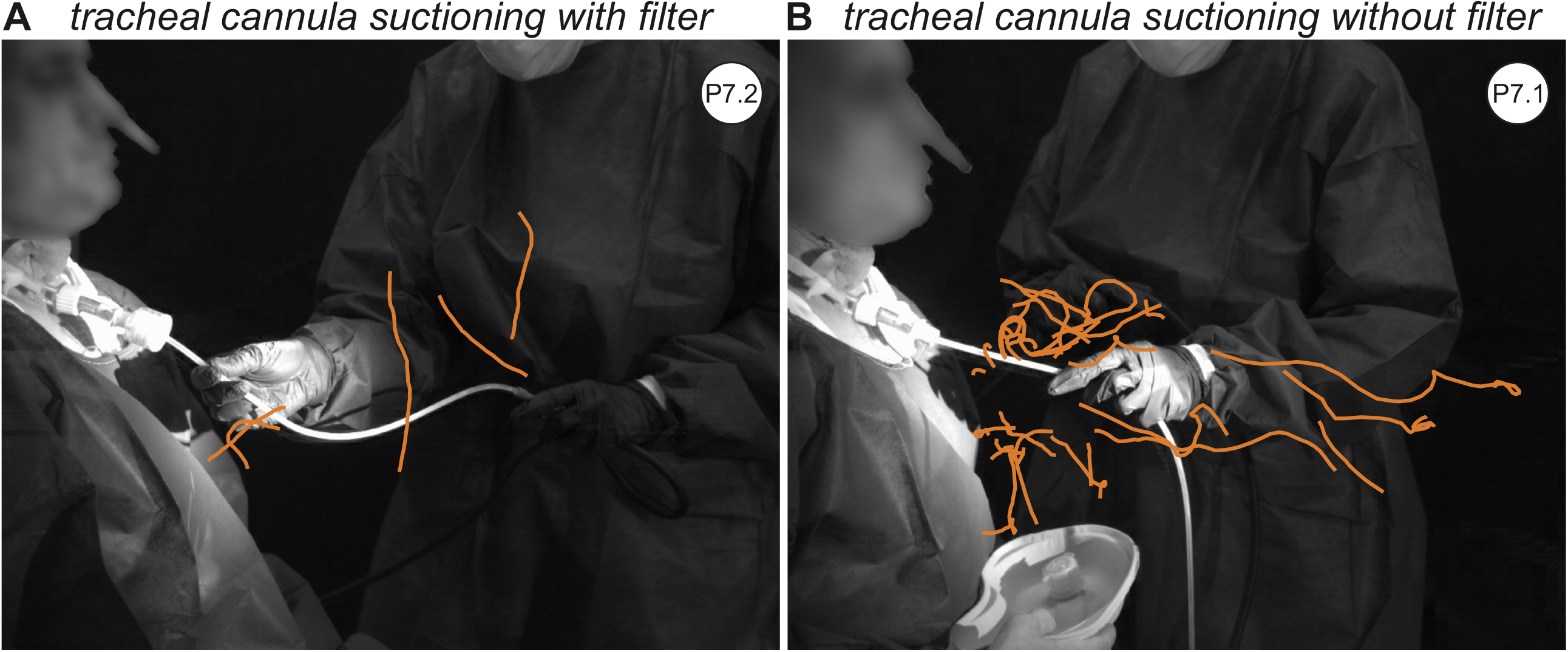
High-speed camera image representing the trajectories of tracheal cannula suction (A) with and (B) without a filter. The orange lines represent the droplet trajectories.

### Speaking and coughing produce more droplets than non-invasive ventilation

Our positive control “stay healthy” as well as coughing showed the highest number (tracking number n=58) of droplets **(video 2)**. In terms of coughing, this is true both for coughing through the mouth without a tracheostomy as well coughing through the tracheostomy opening. Coughing through the mouth demonstrated the fastest velocities with 26.41 m/s.

In contrast, of all non-invasive CPAP procedures and oxygen delivery via a nasal tube only a few scenarios showed droplets. These scenarios included CPAP with PEEP at 10mbar and CPAP with PEEP of 5mbar with a Δp support of 10mbar (paradigms 1 and 2). Low-flow and high-flow of oxygen (2-15l, paradigms 4 and 5) showed no droplets without coughing. However, coughing during nasal oxygen flow generated a large amount of droplets. Here, the amount of droplets was highest during the first cough and decreased for subsequent coughs.

Sampling a SARS-CoV-2 swab according to the WHO guidelines demonstrated a similar effect^18^. During the sampling itself, no droplets could be seen (paradigm 12). However, droplets were detected if the patient spoke or coughed during or after the procedure. During extubation, no coughing and no droplets were visible. At the end of the tracheal tube, gummy secretions were seen (paradigm 11). During nebulization, fine aerosols (manufacturer’s information MMAD 3.6 µm) could be detected using our method. However, aerosols could not be quantified due to their abundance. All results are displayed in **table 1**.

**Table 1:** Conditions, number of recordings, number of recordings where droplets were visible, droplet sizes, trajectory types and computed quantities. The five conditions not provided (i.e. 3, 5, 6, 11, 12) did not show any visible droplets.

| paradigm | # video/<br># with droplets | # tracked | # size | | | # trajRating | | $v_{max}$ (m/s) | | $v_{min}$ (m/s) | | $x_{max}$ (m) | | $dist_{max}$ (m) | | $y_{max}$ (m) | |
| --- | --- | --- | --- | --- | --- | --- | --- | --- | --- | --- | --- | --- | --- | --- | --- | --- | --- |
|  |  |  | s | m | l | t1 | t2 | t1 | t2 | t1 | t2 | t1 | t2 | t1 | t2 | t1 | t2 |
| 1 | 4/1 | 7 | 7 | - | - | - | 7 | - | 0.36 | - | 0.02 | - | 0.21 | - | 0.30 | - | 0.16 |
| 2 | 3/1 | 2 | 2 | - | - | - | 2 | - | 0.11 | - | 0.01 | - | 0.38 | - | 0.21 | - | 0.00 |
| 4 | 5/2 | 40 | 20 | 14 | 6 | 38 | 2 | 26.41 | 4.41 | 0.50 | 0.11 | 0.41 | 0.31 | 0.56 | 0.34 | 0.14 | 0.06 |
| 7.1 | 3/3 | 38 | 34 | - | 4 | 4 | 34 | 0.59 | 7.02 | 0.04 | 0.02 | 0.19 | 0.59 | 0.08 | 0.33 | 0.10 | 0.10 |
| 7.2 | 2/1 | 5 | 5 | - | - | - | 5 | - | 0.33 | - | 0.04 | - | 0.32 | - | 0.18 | - | 0.10 |
| 8 | 2/1 | 5 | 4 | - | 1 | - | 5 | - | 0.45 | - | 0.01 | - | 0.70 | - | 0.13 | - | 0.00 |
| 9 | 2/2 | 15 | 13 | - | 2 | 11 | 4 | 21.69 | 2.35 | 0.12 | 0.01 | 0.73 | 0.28 | 0.73 | 0.06 | 0.08 | 0.01 |
| 10 | 2/2 | 39 | 39 | - | - | - | 39 | - | 1.86 | - | 0.01 | - | 0.39 | - | 0.21 | - | 0.12 |
| 13 | 3/3 | 58 | 45 | 10 | 3 | 53 | 5 | 10.40 | 4.21 | 0.17 | 0.15 | 0.52 | 0.22 | 0.55 | 0.37 | 0.06 | 0.04 |

### FFP3 mask plus surgical mask prevents spread of droplets

We analyzed simultaneously how many droplets originated from the examiner at the same sequences. Although the examiner spoke nearly constantly during the entire procedures, to instruct or calm down the patient, no droplets were seen at any time.

## Discussion

During the COVID-19 pandemic, several studies have emphasized the concept of aerosolized transmission of the virus. SARS-CoV-2 positive aerosols down to a size of 4µm and below were shown to contain contagious viral particles^19^. Around 450.000 health care workers are estimated to have been infected and an appalling number have died^20^. As a result, several routine airway management procedures are currently being adapted or avoided in SARS-CoV-2 positive patients. This is the first study to evaluate common airway management procedures performed in real-life patients in order to characterize droplet patterns.

We applied high-speed imaging to visualize droplets in the region of interest (entire dispersion of droplets). For our study, we performed the airway management procedures as realistically as possible on patients. We focused on larger respiratory droplets rather than on airborne aerosols (smaller than 5 µm) due to the suspected higher viral load^15,16^. In our results, two different trajectory patterns of the droplets emerged which we categorized as type 1 and 2. All droplet sizes are found in both trajectory types: type 1 (i.e. classic ballistic trajectory) and type 2 (i.e. driven by convective forces of the air resulting from the air management procedure itself). Due to the low mass of the droplets, convective lifting forces balance with their gravitational forces resulting in a floating behavior similar to much smaller aerosols. This results in a non-predictive, complex, trajectory pattern, alternating accelerations and decelerations capable of reaching the provider’s face. To the best of our knowledge, this behavior has not been described for such large droplets during these air management procedures in patients before. We could show that the droplets described in type 2 remained in the air for a minimum of 7 seconds. With an average human being roughly breathing every 5 seconds (12/min), we suggest that the droplets persist long enough for another person to inhale. We hypothesize that by remaining longer in the air and recirculating non-predictively, type 2 droplets are more dangerous for disease transmission than type 1 droplets. Importantly, our type 2 pattern is similar to previously described aerodynamic patterns of fine aerosols (< 5 µm) although droplets were significantly larger. Whether these larger droplets are more contagious than the fine particle aerosols with the same aerodynamic pattern due to a potential higher viral load remains to be verified in virological experiments.

Our results also confirm that the usage of a filter significantly reduced the amount of droplets in comparison to a non-occluded tracheal cannula during tracheal cannula suctioning. Additionally, we could not detect any droplets originating from the examiner who was wearing the combination of a FFP3 and a surgical mask. The same examiner, however, had shown a droplet formation during the positive control “stay healthy” without a mask. Various international guidelines recommend the use of a viral filter e.g. in combination with a heat-moisture exchanger during intubation and extubation^21–24^. A previous study of our group showed significant reduction of droplets during tracheal cannula procedures using a filter^25^. However, the filter did not avoid droplet spread completely which leaves health care workers still at risk. Additionally, the use of combination of a FFP3 and a surgical mask as source control is generally assumed to be one of the most important measures to prevent airborne virus transmission^26^. Our results confirm and strengthen these recommendations.

Furthermore, our results show that speaking and coughing produced the highest amount of droplets with the highest velocities. In previous research^27^, the sentence “stay healthy” was shown to produce speech associated droplets. Our study confirmed that coughing and speaking produced the largest amount of droplets over all conditions. Coughing demonstrated the highest velocity measurements and the furthest distance travelled. Our velocity during coughing was even higher than reported in previous studies^28^. With respect to particle numbers, speaking and coughing produced significant more droplets than routine airway management procedures including CPAP non-invasive ventilation, high-flow oxygen administration using a nasal cannula and extubation. As recommended in literature^7,9,29^, in a highly sedated, ventilated patient with a blocked cuff at either breathing tube or tracheal cannula and an intercalated viral filter, droplet formation is minimal. Our results show that even non-invasive ventilation in a breathing patient shows minimal droplet concentration. However, the awake patient that is able to cough and speak appears to be the most dangerous. As a patient on non-invasive CPAP or nasal cannula ventilation is awake, the patient is potentially able to cough and speak which represents a potential risk. These findings underlie the unconditional need of personal protective equipment (PPE). Summarizing, our results show that aerodynamic measurements during airway management procedures are able to provide valuable information for the safety of the medical personnel and for the patients.

## Conclusions

Large respiratory droplet patterns generated during airway management procedures follow two trajectories, one ballistic, and one approximating that of smaller airborne particles following a random convective pattern. Speaking and coughing produced both a larger amount and higher velocity droplets as compared to the investigated airway management procedures. Facial masks significantly reduced droplet dissemination as did the use of a tracheal cannula filter.

## Data Availability

Data openly available in a public repository that issues datasets with DOI

## Abbreviations

ARDS: acute respiratory distress syndrome
COVID-19: coronavirus disease 2019
CPAP: continuous positive airway pressure
FFP: filtering face piece
FPS: frames per second
LED: light-emitting diode
MMAD: mass median aerodynamic diameter
PEEP: positive endexpiratory pressure
PPE: personal protective equipment
Δp support: pressure support ventilation
SARS-CoV-2: severe acute respiratory syndrome coronavirus 2
WHO: World Health Organization

## Figures and movies

**Video 1:** High-speed recording of a person coughing through a tracheostomy opening without a tracheal cannula.

**Video 2:** High-speed recording of a person saying “stay healthy”

